# The relationship between level of education and disease onset and progression in Spinocerebellar Ataxia types 1, 2, 3, and 6

**DOI:** 10.64898/2025.12.10.25342000

**Authors:** Collin J Anderson, Daria Nesterovich Anderson, Ashley Kucharski, CRC-SCA Consortium, Vikram Shakkottai, Sheng-Han Kuo, Liana S. Rosenthal

## Abstract

**Background:** Socioeconomic factors such as educational attainment are associated with altered progression in numerous neurodegenerative disorders, but this relationship has not been fully explored in the context of spinocerebellar ataxias (SCAs). In previous work from several single-center studies, educational attainment was associated with modified onset or increased progression in SCA3 or SCA6. The relationship between such socioeconomic factors and onset or progression in SCAs has not previously been reported using data from studies conducted across multiple sites.

**Methods:** We evaluated data from the North America-based Clinical Research Consortium for the Study of Cerebellar Ataxias collected from 18 sites. We evaluated genetic, onset, severity, and self-reported educational data from patients with confirmed diagnoses of SCA types 1, 2, 3, or 6 and built linear mixed-effects models to evaluate the association between the level of education against disease onset and severity at a given timepoint.

**Results:** In this cohort, we found that increased educational attainment was not associated with age of onset in any of the evaluated SCAs. However, in the context of severity at a given time point, increased education was strongly linked to reduced severity specifically in SCA3, but not in SCA types 1, 2, or 6. Results were consistent across several forms of robustness analysis and validation.

**Conclusions:** In a North American cohort, severity of SCA3, but not SCA1, SCA2, or SCA6 was found to be strongly inversely correlated with level of educational attainment. Further study in other cohorts would be useful.

## Introduction

Spinocerebellar ataxias (SCAs) are a group of several dozen autosomal dominant hereditary ataxias. Numerous SCAs are caused by polyglutamine repeat (CAG) expansions, including SCA types 1, 2, 3, and 6, which are caused by expansions within the ATXN1, ATXN2, ATXN3, and CACNA1A genes, respectively^1–4^. Each of these SCAs features an inverse relationship between age of onset and expanded allele repeat length, with repeat expansion predicting about 40-70% of variability in age of onset across these SCAs^5–8^. In contrast to the clear link between CAG repeat expansion length and age of onset, its effect on symptom progression of SCAs is less clear. In SCA types 1, 2, and 3, it has often been shown that increased CAG repeat expansion is linked to faster progression^9–11^, but this finding has not been replicated in all studies^12^. Further, in SCA6, CAG repeat expansion length does not appear to be linked to the rate of progression.^13^ Thus, it may be that while CAG repeat expansion length is strongly anticorrelated to age of onset in each of SCA types 1, 2, 3, and 6, it is at most only modestly tied to disease progression in a subset of SCAs.

Better ability to predict age of onset and disease progression in SCAs would be valuable for numerous reasons, including but not limited to improved clinical management, more personalized prognoses, improved genetic counseling, optimized clinical trials, biomarker development, and improved resource allocation. Socioeconomic factors play an important role in the onset and progression of numerous neurodegenerative disorders, such as Parkinsonism^14^, various dementias^15^, and other CAG repeat expansion disorders such as Huntington’s disease^16^. Given that level of education is correlated with numerous other socioeconomic factors, it is a reasonable proxy for socioeconomic status when evaluating factors related to disease onset and progression.

A recent single-center study found that level of education was inversely correlated with both age of onset and progression in a cohort of SCA3 patients, suggesting that increased education might be associated with earlier onset but slower progression in SCA3^17^. Further, another recent single-center study found that increased education might be associated with reduced severity and slower progression in a cohort of SCA6 patients^13^. As a follow up, in this study, we aimed to expand this analysis to a broader North American cohort and several additional SCAs through use of data from the Clinical Research Consortium for the Study of Cerebellar Ataxia (CRC-SCA)^18^. Thus, we sought to address two primary questions: first, do the above-mentioned findings in the context of SCA3 or SCA6 hold in this larger cohort, and second, is educational attainment likewise associated with age of onset and/or severity in other CAG repeat expansion SCAs?

## Methods

### General overview

Data were obtained from the Clinical Research Consortium for the Study of Cerebellar Ataxia (CRC-SCA), co-chaired by authors LSR, VGS, and SHK, in February 2025. From the larger dataset, we systematically applied our inclusion criteria (discussed in *Subjects* below) to isolate data for subjects diagnosed with SCA types 1, 2, 3, and 6. Across several separate linear mixed-effects models, we first evaluated the independent contributions of expanded allele repeat length and education on age of onset, followed by the independent contribution of each of expanded allele repeat length, age of onset, duration of disease, and educational attainment on Scale for Assessment and Rating of Ataxia (SARA) score. Next, we generated predicted SARA scores via a linear mixed-effects model with inputs of expanded allele repeat length, and the duration of disease. We compared predicted SARA scores to actual recorded SARA scores to calculate a residual, then evaluated the relationship between residuals and years of education to determine whether educational attainment correlated with SARA score and thus might influence severity at a given amount of time post onset (and by proxy, progression). Finally, when significant results were found, we performed multiple analyses of robustness and validation (discussed in *Statistics* below).

### Ethics

CRC-SCA includes patient data from 18 clinical sites across Minnesota, California, Washington, Utah, Texas, Illinois, Michigan, Florida, Georgia, Maryland, Pennsylvania, New York, and Massachusetts in the US, and Québec in Canada. Data were collected under a uniform protocol approved by each institution’s IRB, with written informed consent from each patient. Data were then made available to researchers in a deidentified fashion. While authors at locations other than the University of Sydney already have approved IRBs for collection and analysis of this work at their respective institutions, the authors at University of Sydney received a written exemption from the University of Sydney Human Ethics Committee to complete these studies without a corresponding human ethics protocol based on the Australian National Health and Medical Research Council’s definition of negligible risk in the National Statement on Ethical Conduct in Human Research. This was based on meeting the criteria of “negligible risk research” and involving “the use of existing collections of data or records that contain only non-identifiable data about human beings.”

### Subjects

We included patients who had a genetically confirmed case of SCA type 1, 2, 3, or 6, in addition to a reported expanded allele repeat length, and a reported age of onset. When the age of onset was not directly reported in the appropriate column, we used the first reported age/year of first SCA symptom onset instead, assuming age of onset is based on the earliest symptomatic onset. We also included one or multiple SARA scores of symptom severity for a large subset of patients when a motor evaluation was completed, and for which the subject’s age at which the evaluation was also recorded. Analyses of education included a smaller subset of subjects who reported educational attainment. General group analyses included patients who met the minimum criteria for the analysis question, whether or not educational attainment was self-reported or SARA score was evaluated. Patients were automatically excluded prior to any analyses when obvious and unresolvable data entry errors were observed, such as if a non-zero SARA score was reported at an earlier timepoint than the reported age of onset.

In total, our dataset included 39 SCA1 patients, of whom 28 reported educational attainment and 27 reported educational attainment and had severity data at one or more time points. There were 80 SCA2 patients, of whom 61 reported educational attainment and 54 reported educational attainment and had severity data at one or more time points. Our largest cohort sample with our minimum inclusion criteria was 123 SCA3 patients, of whom 89 reported educational attainment and 83 reported educational attainment and had severity data at one or more time points. Finally, there were 61 SCA6 patients, of whom 48 reported educational attainment and 43 reported educational attainment and had severity data at one or more time points (Table 1). Ethnicity and race data are reported in supplemental materials (Supplemental Table 1).

**Table 1:** Demographic and clinical data are shown for each SCA cohort. This includes gender (percent female) as well as mean and standard deviation of age of onset, expanded allele length, and normal allele length from all participants. Reported years of education are averaged from only those who provided educational data, and disease duration and SARA score are from only those who both provided education and had recorded SARA scores at a given timepoint.

| SCA type | Number of patients | Number of patients with educational data | Number of patients with educational and SARA data | % Fem | Age | Expanded allele repeat length | Normal allele repeat length | Years of education | Disease duration (years) | SARA score |
| --- | --- | --- | --- | --- | --- | --- | --- | --- | --- | --- |
| SCA1 | 39.0 | 28.0 | 27.0 | 56.4 | 41.1 ± 12.6 | 46.7 ± 4.7 | 29.6 ± 1.9 | 15.3 ± 2.4 | 6.1 ± 6.8 | 12.1 ± 8.1 |
| SCA2 | 80.0 | 61.0 | 54.0 | 67.5 | 38.6 ± 14.0 | 40.8 ± 3.5 | 22.2 ± 0.9 | 15.0 ± 3.0 | 9.5 ± 9.2 | 13.4 ± 7.0 |
| SCA3 | 123.0 | 89.0 | 83.0 | 59.3 | 42.1 ± 13.0 | 71.3 ± 4.2 | 22.6 ± 6.4 | 15.4 ± 2.5 | 7.8 ± 7.5 | 11.2 ± 7.4 |
| SCA6 | 61.0 | 48.0 | 43.0 | 50.8 | 55.1 ± 10.0 | 22.3 ± 0.8 | 12.4 ± 1.2 | 15.4 ± 2.5 | 8.2 ± 8.0 | 11.0 ± 6.8 |

### Assumptions made surrounding years of education

Participants self-reported highest level of education as one of the following options: <12 years, high school or GED, some college, associate’s degree, bachelor’s degree, master’s degree, or doctorate degree. To codify these discrete categories into numerical values, we made the following standard assumptions: we assigned a numerical value of 10 years of education to all individuals who self-reported <12 years of education. High school or GED recipients were assigned 12 years of education. As most university dropouts complete less than 2 years of their university degree, we assigned 13 years of education to all individuals who reported some college. Further, those who reported completing an associate’s degree were assigned 14 years of education, those who reported completing a bachelor’s degree were assigned 16 years of education, and those who reported completing a master’s degree were assigned 18 years of education. Finally, while doctorate degrees can be variable in duration, those who reported a doctorate degree were assigned 20 years of education.

### Linear mixed-effects models

For each of SCA types 1, 2, 3, and 6, we generated several linear mixed-effects models (LMMs) within the Python environment (version 3.12). Prior to any linear mixed-effect model generation, we carried out regression analyses to confirm that greater repeat expansions were associated with earlier symptom onset. Next, for each SCA, we generated LMMs to evaluate the impact of educational attainment on age of onset independently of expanded allele repeat length.

We next generated several LMMs to evaluate the impact of educational attainment on severity. First, for each SCA, we generated a linear mixed-effects model that evaluated the impact of educational attainment independently from age of onset, expanded allele repeat length, and duration of disease. Next, through a leave-one-out approach and using age of onset, expanded allele repeat length, and duration of disease as variables, we iteratively removed each patient and predicted their SARA score from a linear mixed-effects model comprising data from all remaining patients within that SCA type for each subject’s datapoint. Educational attainment was not included in this model. To explore this, we calculated the marginal residual in this predicted SARA score compared to the observed SARA score and then correlated this residual value (i.e. the difference between prediction and observation) to educational attainment. Thus, for n patients with any SCA type, we generated n LMMs fit using data from n-1 patients, each tested on the nth patient. If a subject had multiple timepoints for recorded SARA score, the same LMM was used to predict the SARA score for that patient for each timepoint. Notably, for each LMM in the context of SARA score, this calculation occurs at a given time point — reduced severity at a specific timepoint since onset should be interpreted as slower progression.

Following the above, we carried out several robustness-related analyses in the context of significant results surrounding severity-related outcomes. First, given a collinearity between age of onset and expanded allele repeat length in the above models, we repeated the prediction of SARA score based solely on age of onset and duration of disease, in the absence of expanded allele repeat length or education. While such models were less accurate in predicting SARA than those including expanded allele repeat length, they enabled a similar analysis of education in the absence of violated collinearity assumptions (between expanded allele repeat length and age of onset). Second, while the above analyses were carried out via a leave-one-out approach, we performed confirmatory secondary analyses of significant findings through an alternate bootstrap approach. In this approach, for a given SCA, with n patients in the dataset, we sampled and built a model based on n patients with replacement, and from the resulting model, we predicted SARA score from the remaining unchosen patients. We repeated this process 10,000 times. We found the mean of all SARA predictions for each timepoint for each patient and then repeated residual (predicted minus observed) analyses as described above. Finally, for most SCAs, we noticed that models tended to consistently underpredict high observed SARA scores. In the context of SCA3, we noticed a skew, which was statistically confirmed, that those with higher SARA scores (particularly 20+) has with less education. Thus, given significant primary findings in the context of SCA3, we repeated analyses for only SARA scores under 20 to ensure robustness.

### Statistics

Using the statsmodels package in Python, statistics were reported direction from the ols or mixedlm functions for all regressions, LMMs, leave-one-out, and bootstrap approaches. Significance tests for LMM analyses were performed within the Python package via a Wald test, including for analyses of the model residual versus education, which still employed LMMs based on multiple timepoints from numerous subjects. In the case of expanded allele repeat length versus age of onset, we note that, given only two variables, the Wald test is identical to a Pearson correlation test. In each LMM, we evaluated model conditions in the context of symmetry of residuals (skew), peakedness or tail heaviness of distributions (kurtosis), normality (omnibus and Jarque-Bera, both based on skew and kurtosis), autocorrelation of residuals (Durbin-Watson), and collinearity (condition number). When any assumption failed (occurred only in terms of collinearity of age of onset and expanded allele repeat length), additional models were developed as described above to confirm robustness.

## Results

### Subject demographics

Demographic data for all subjects for each SCA type are shown in Table 1. Three separate patient counts are given. First presented are the numbers of patients evaluated in the study with the given SCA type, with a requirement of only age of onset and expanded allele repeat length data; second, the numbers of patients with age of onset, expanded allele repeat length, and self-reported level of education; and finally, the numbers of patients with each of age of onset, expanded allele repeat length, self-reported level of education, and at least one SARA score at a given age. Additionally reported are subjects reported as female and average (and standard deviation, when relevant) age of onset, expanded allele repeat length, disease duration, SARA score, self-reported level of education, gender, and race.

### Expanded allele repeat length is negatively associated with age of onset

For each SCA reported, we evaluated the relationship between expanded allele repeat length and age of onset (Figure 1A). In each case, we found a strong, significant negative relationship between repeat length and age of onset, matching findings in numerous previous reports. Coefficients and p-values for Figure 1A and all other results except those in supplemental figures are reported in Table 2.

**Figure 1:**
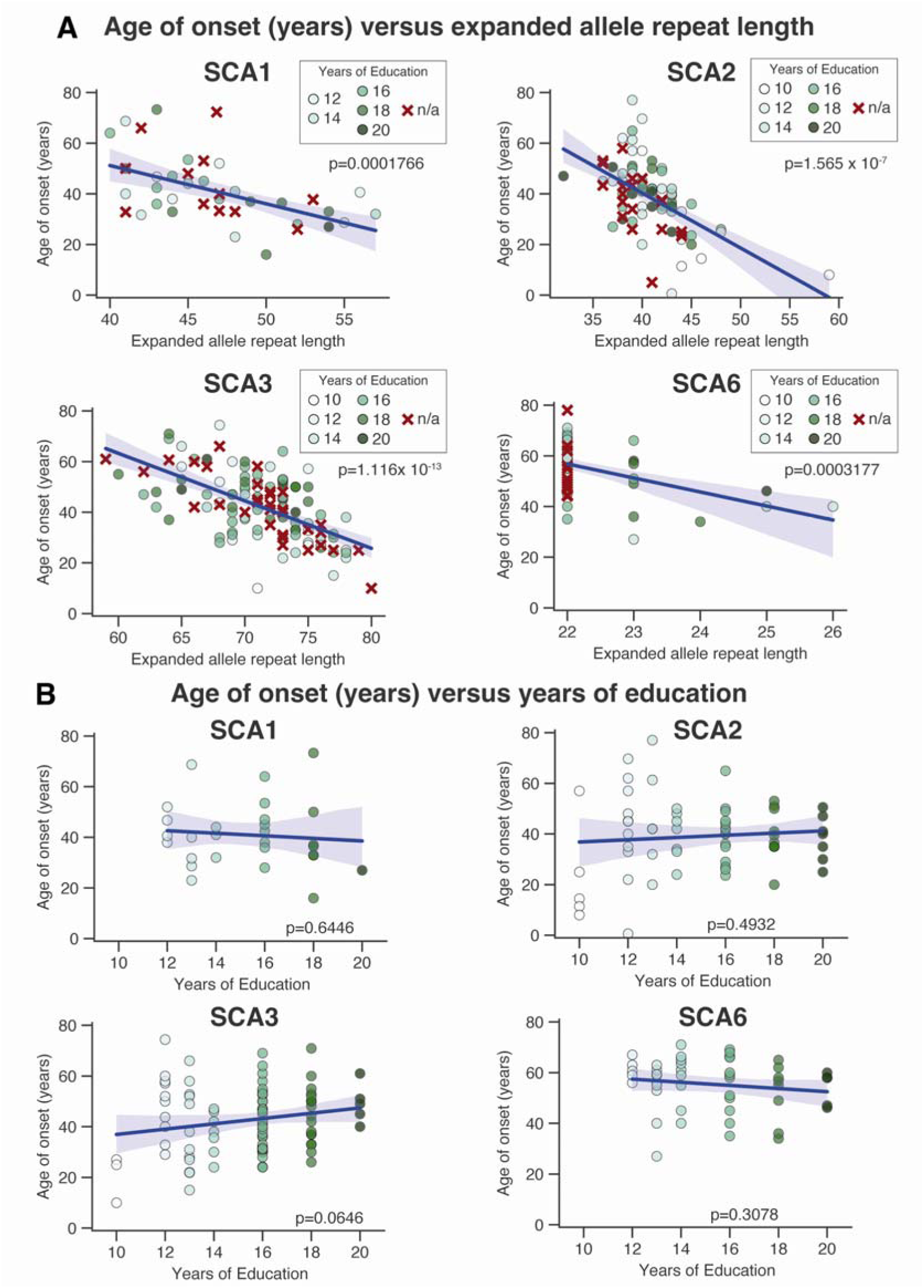
**A.** For each of SCA type 1, 2, 3, and 6, increased expanded allele repeat length was strongly associated with earlier onset, as expected. Note that color of dot corresponds to years of education, while patients without reported educational data are shown with a red x. **B.** Educational attainment was not associated with age of onset in any SCA type in this cohort. Note that beyond the simplified visualized regression methods shown, we evaluated whether educational attainment underlay age of onset in a LMM, and the trend shown here in SCA3 weakened when accounting for expanded allele repeat length.

**Table 2:**
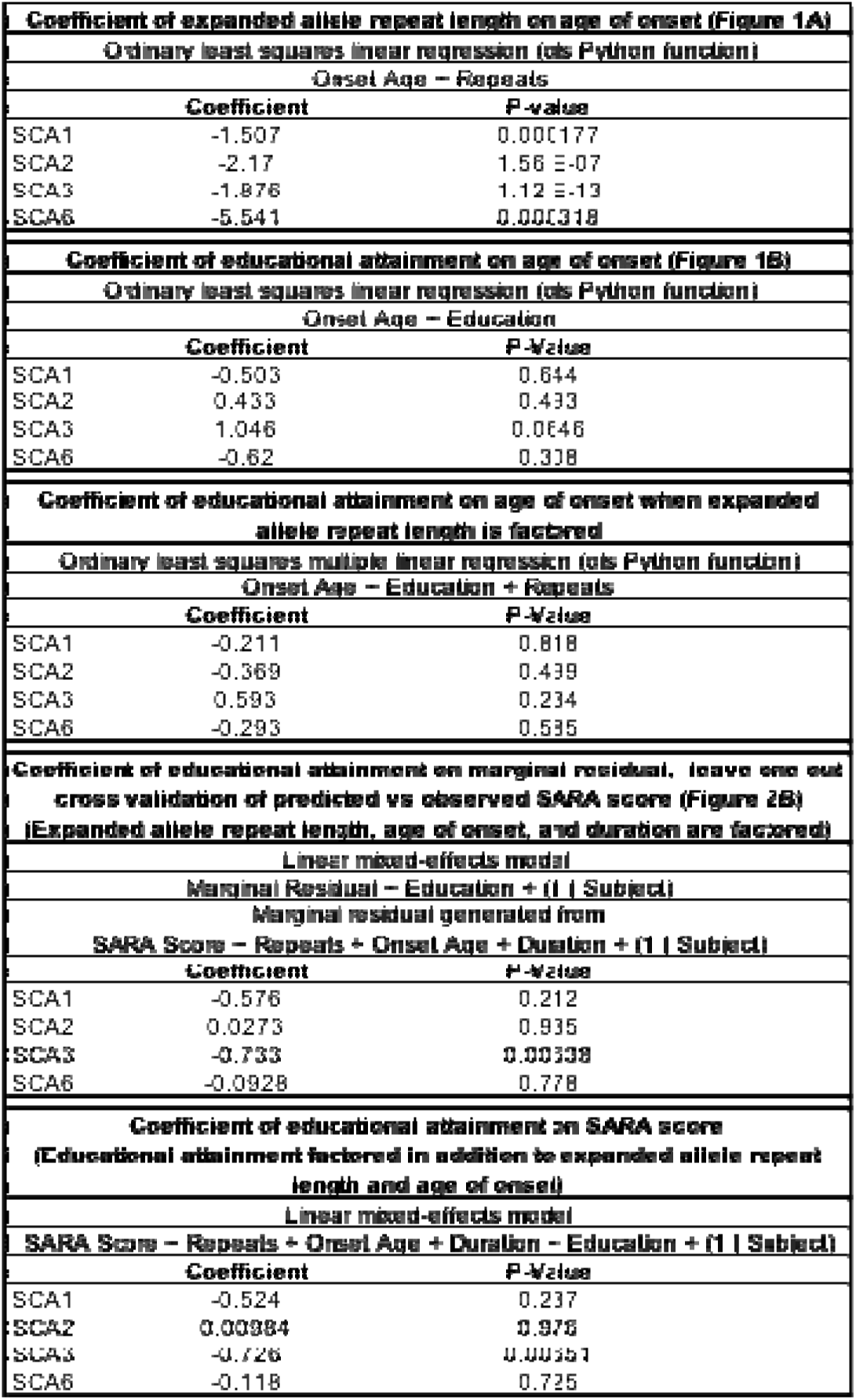
Coefficients (slopes) and p-values are shown for all analyses, with the exception of robustness analyses, which are provided in supplemental materials.

### Educational attainment was not found to modify age of onset in this cohort

We generated LMMs for each reported SCA to evaluate the independent effects of expanded allele repeat length and educational attainment on age of onset. For each SCA, we found insignificant relationships between educational attainment and age of onset (statistics results presented in Table 2). In Figure 1B, for the ease of visualization, we show simplified regression relationships between education and age of onset, and we specifically make note that the strong trend in SCA3 shown in this figure was weakened substantially when the effect of expanded allele repeat length was accounted for in the above-mentioned LMM. Further, we note that trends in SCA3 did not match previous reports that greater educational attainment was associated with earlier age of onset^17^.

### Greater educational attainment was found to be correlated with reduced severity in SCA3, but not in other SCAs in this cohort

For each SCA type, we generated LMMs that predicted SARA score for a given timepoint for n patients based on patient identity, age of onset, expanded allele repeat length, and disease duration. Model predictions versus observed SARA values are shown in Figure 2A. For each SCA, we found marginal residuals—predicted minus observed SARA—and evaluated the relationship between marginal residual and educational attainment (Figure 2B). For each of SCA types 1, 2, and 6, no relationship was found between educational attainment and marginal residual, meaning that educational attainment did not account for the discrepancy from observed to predicted. However, for SCA3, increased educational attainment was negatively correlated with the marginal residual, meaning that increased educational attainment was associated with reduced severity than predicted in this cohort. Given that progression is change in severity over a duration of time, reduced severity at a given time should be interpreted as slower progression.

**Figure 2:**
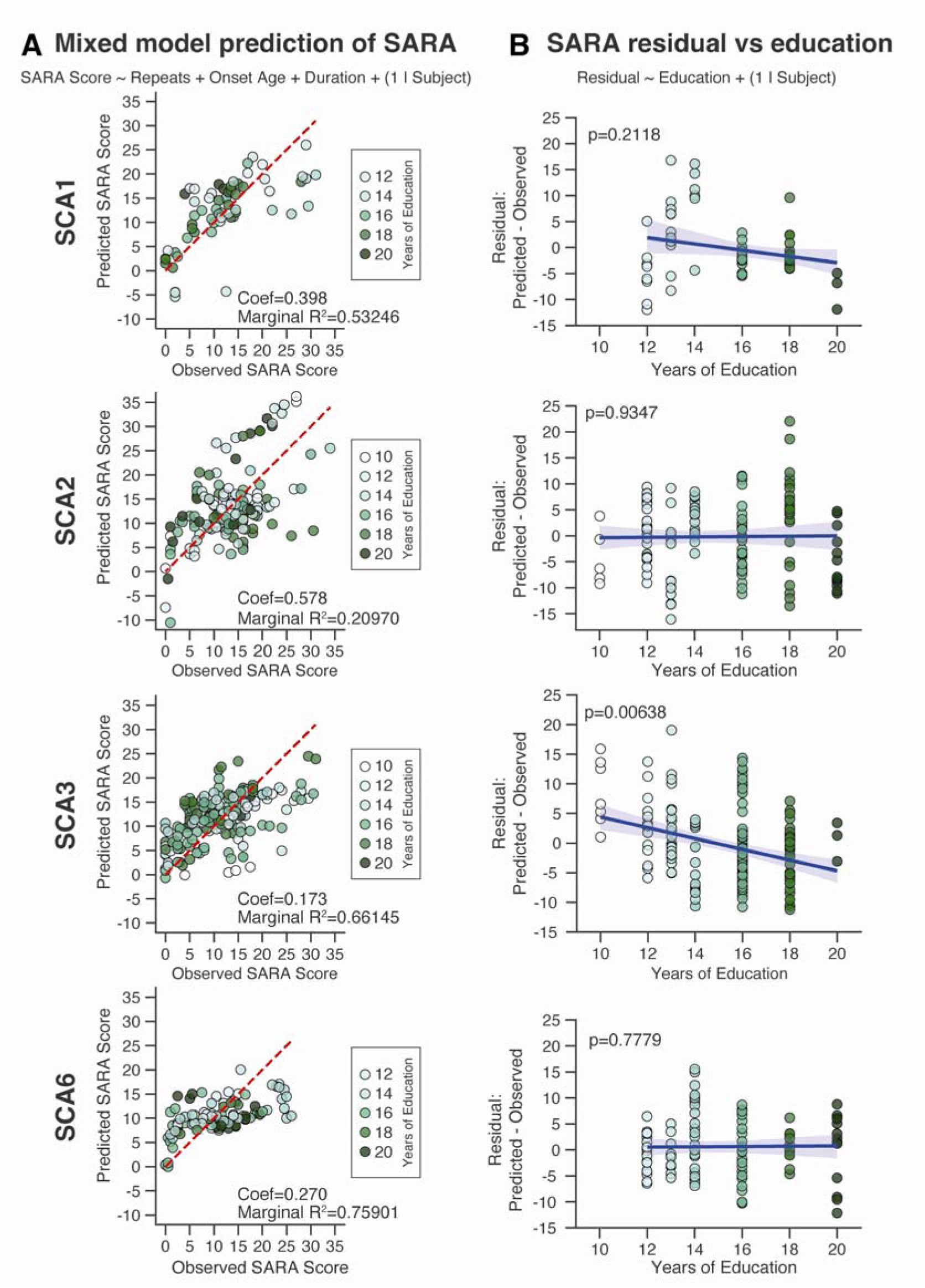
**A.** LMMs were generated to predict SARA score specific to a subject at a given age from expanded allele repeat length, age of onset, and duration. **B.** Educational attainment was found to be strongly negatively correlated with error in model prediction (residual) in SCA3, but not in the other SCAs tested. Thus, in this cohort, higher educational attainment is associated with slower progression in SCA3 but not in SCA types 1, 2, or 6.

Further, we generated LMMs for each reported SCA to with the following input variables: patient identity, age of onset, expanded allele repeat length, duration, and education, in order to evaluate the independent effect of education on SARA. In alignment with the above, education did not significantly contribute to SARA for SCA types 1, 2, or 6 in this cohort, but it did for SCA3.

### The relationship between educational attainment and severity held up to multiple forms of robustness testing

We performed several robustness analyses to further validate our findings that higher education is associated with reduced disease severity at specific time points (and by proxy, slower progression) in SCA 3 but not SCA 1, 2, or 6 in this dataset. First, we found high collinearity between age of onset and expanded allele repeat length within each SCA type. Thus, to pass model assumptions, we repeated all experiments reported above in Figure 2 in the context of a LMM with solely onset age and duration as inputs, but without expanded allele repeat length. While numerical outcomes differed, conclusions matched those described above (Supplemental Figure 1). Second, while the findings in Figure 2 were reported based on a leave-one-out approach, we validated further using a bootstrap confidence assessment with replacement, finding similarly significant results. Finally, evaluating SCA3 results, we observed a potential skew in the dataset; specifically, upon evaluating, we found that SARA scores >20 occurred more in patients with less educational attainment. Given that the LMMs tended to underpredict SARA when observed SARA was high, we generated a new SCA3 LMM specifically for only SARA scores <20 to ensure that results were not driven by this skew. In this final form of robustness testing, we again found that conclusions in the context of SCA3 held, namely that increased education was associated with reduced severity (Supplemental Figure 2). Statistics for results related to supplemental figures are shown in Supplemental Table 2.

## Discussion

In this work, we aimed to determine whether educational attainment was associated with modified age of onset or severity in spinocerebellar ataxia types 1, 2, 3, and 6 in a North American patient cohort sourced from the Clinical Research Consortium for the Study of Cerebellar Ataxia, supported by the National Ataxia Foundation. We started by evaluating the association between expanded allele repeat length and age of onset for each, finding robust, significant correlations for each SCA, in line with numerous previous reports. Next, and similarly unsurprisingly, we found that increased expanded allele repeat length, earlier age of onset, and longer disease duration were significantly associated with higher SARA values at a given time point in each SCA type. Following these confirmatory analyses, we evaluated the association between educational attainment and each of age of onset and severity at given time points. In this cohort, we did not find an association between education and age of onset. We found a significant relationship between education and reduced severity in SCA3, but not in SCA types 1, 2, and 6. Furthermore, these findings in the context of SCA3 held up to multiple forms of analysis in the context of robustness. Thus, in this cohort, we conclude that educational attainment is associated with reduced severity—and by proxy, slower progression—in SCA3, but not in SCA1, SCA2, or SCA6.

It is notable to consider our results in the context of previous reports. In the context of SCA3, previous evaluation of a Johns Hopkins Ataxia Center patient cohort showed that increased educational attainment was associated with slower progression of SARA score^17^, which closely aligns with our reported findings. However, in that cohort, increased education was significantly associated with earlier age of onset, whereas in our study, we failed to find a significant association, and instead, we found a trend in the opposite direction of these previously reported findings. Further, we failed to find agreement with a recent study that showed that higher educational attainment was associated with decreased SCA6 severity and slower disease progression in a SCA6 Network at the University of Chicago SCA6 cohort^13^. Notably, this University of Chicago study featured a larger sample size; however, our findings show no trend in the context of SCA6, and thus, differences aren’t likely sample size dependent. However, patients in our study were, on average, only 8 years post onset, while those in the University of Chicago study averaged 20 years post onset. Thus, with more time, perhaps our cohort would match the University of Chicago cohort.

It is important to consider the effect size we found for our primary results in SCA3. As shown in Table 2, for SCA3 subjects, the coefficient of educational attainment SARA is −0.726. Thus, at the average timepoint in our cohort (7.8 years post onset in SCA3), each additional year of education is linked to, on average, a 0.726 point reduction on the SARA scale. This corresponds to 0.1 points on SARA per year difference in progression for each year of education. This would yield an average of a 7.26-point difference in SARA between individual that didn’t complete high school vs individuals with a doctorate degree in 7.8 years or nearly a 1-point difference per year in this comparison.

It is worthwhile to reflect on why we found a strong association between educational attainment and severity in SCA3 but not SCA types 1, 2, and 6. First, it may be relevant to consider the specific patterns of neurodegeneration across each SCA type. Specifically, SCA3 patients may have more or earlier cerebral cortical decline than SCA 1, 2, or 6 patients^19,20^. Neurodegenerative disorders for which increased educational attainment has been most commonly linked to slower progression are often, but not always, those with substantial cortical involvement ^21–23^, directly relevant to the concept of cognitive reserve^24^. In contrast, in Parkinsonism, which has minimal early cortical involvement, educational attainment is linked to reduced motor burden, although this has been hypothesized to be based on extranigral protective effects.^25^ Based on all of the above, it may stand to reason that the association between increased educational attainment and slower progression in SCA3 could be based on the greater degree of extracerebellar involvement in SCA3. However, this would not explain the decreased SCA6 severity and slower disease progression from the University of Chicago SCA6 cohort with greater education^13^, given SCA6’s minimal extracerebellar involvement.^26^ Second, and more simply, it is possible that the nature of cognitive and motor impairments in SCA1 and SCA2 may be less amenable to modification via educational attainment. Indeed, in SCA1 and SCA2, repeat length has a much stronger contribution to progression than it does in SCA3 of SCA6^27^, and thus, there may be more ability to modify progression through educational attainment in SCA3 or SCA6.

Several limitations should be addressed. It is important to note that this study was limited to a North American cohort, and educational attainment may be either more or less relevant as a socioeconomic factor in other regions of the world. It is also important to consider that there may be interactions between how ethnicity and educational attainment are associated with progression in some populations. For example, SCA3 is more than an order of magnitude more common in Aboriginal communities in the Groote Eyelandt Archipelago of Australia than in any other community worldwide^28^, and findings in such a cohort in the context of education as a socioeconomic factor might not necessarily mirror those reported in this work. Second, an obvious question emerges as to whether an absence of findings in SCA1, SCA2, and SCA6 in the context of education and severity, was simply dependent on lack of power. In the context of SCA2 and SCA6, the near-zero coefficients despite robust sample sizes (n=54, n=43, respectively) suggests a true absence of a relationship. SCA1 shows a potential trend (p=.212) with a smaller cohort (n=27), and thus it may be reasonable to speculate that this study was simply underpowered in the context of SCA1. However, we note that the trend weakened in both robustness analyses that included SCA1 (p=.342, .232 for the two analyses), and thus, there may simply not be an association between educational attainment and severity in this this cohort of SCA1 patients. However, it would be impossible to draw a definitive conclusion without more data. Further, it’s notable that numerous additional factors contribute to SCA onset and progression, and their inclusion in LMMs may power improved analysis as to specific, independent effects of educational attainment. However, data were often limited, and thus, we limited our model to those variables that were consistently available, increasing our sample size. Regardless, if more data were available, future analyses could be expanded, say, to include additional genetic data for genetic modifiers.

In this work, we evaluated the impact of educational attainment on age of onset and severity or progression in North American patients with SCA types 1, 2, 3, or 6. In contrast with prior work, we did not find a significant impact of education on age of onset. However, we did find that educational attainment was strongly linked to reduced severity or slower progression specifically in SCA3. This work fits into a larger social determinant of health model of disease progression and future work should incorporate additional components of socioeconomic status and access to care. Future work should also explore these results in additional datasets, such as the European Integrated Project on Spinocerebellar Ataxias (EUROSCA), would be useful in generating increased ability to predict onset and progression across SCA patients.

## Data Availability

All data produced in the present study are available upon reasonable request to the authors.

## Supplemental Materials

**Supplemental Table 1:** Ethnic and racial data are shown as a percentage of all patients within the specific SCA.

|  | SCA1 | SCA2 | SCA3 | SCA6 |
| --- | --- | --- | --- | --- |
| **Ethnicity** |  |  |  |  |
| Not Hispanic, Latino, or Spanish | 94.87 | 88.75 | 92.68 | 91.80 |
| Hispanic, Latino, or Spanish orig | 2.56 | 7.50 | 6.50 | 4.92 |
| N/A Ethnicity | 2.56 | 3.75 | 0.81 | 3.28 |
| **Race** |  |  |  |  |
| White | 66.67 | 62.50 | 59.35 | 83.61 |
| Black or African American | 12.82 | 17.50 | 27.64 | 0.00 |
| Asian | 12.82 | 12.50 | 8.94 | 11.48 |
| Asian, White | 2.56 | 0.00 | 2.44 | 0.00 |
| Native HI or Pacific Islander | 0.00 | 0.00 | 0.81 | 3.28 |
| N/A Race | 5.13 | 7.50 | 0.81 | 1.64 |

**Supplemental Figure 1:**
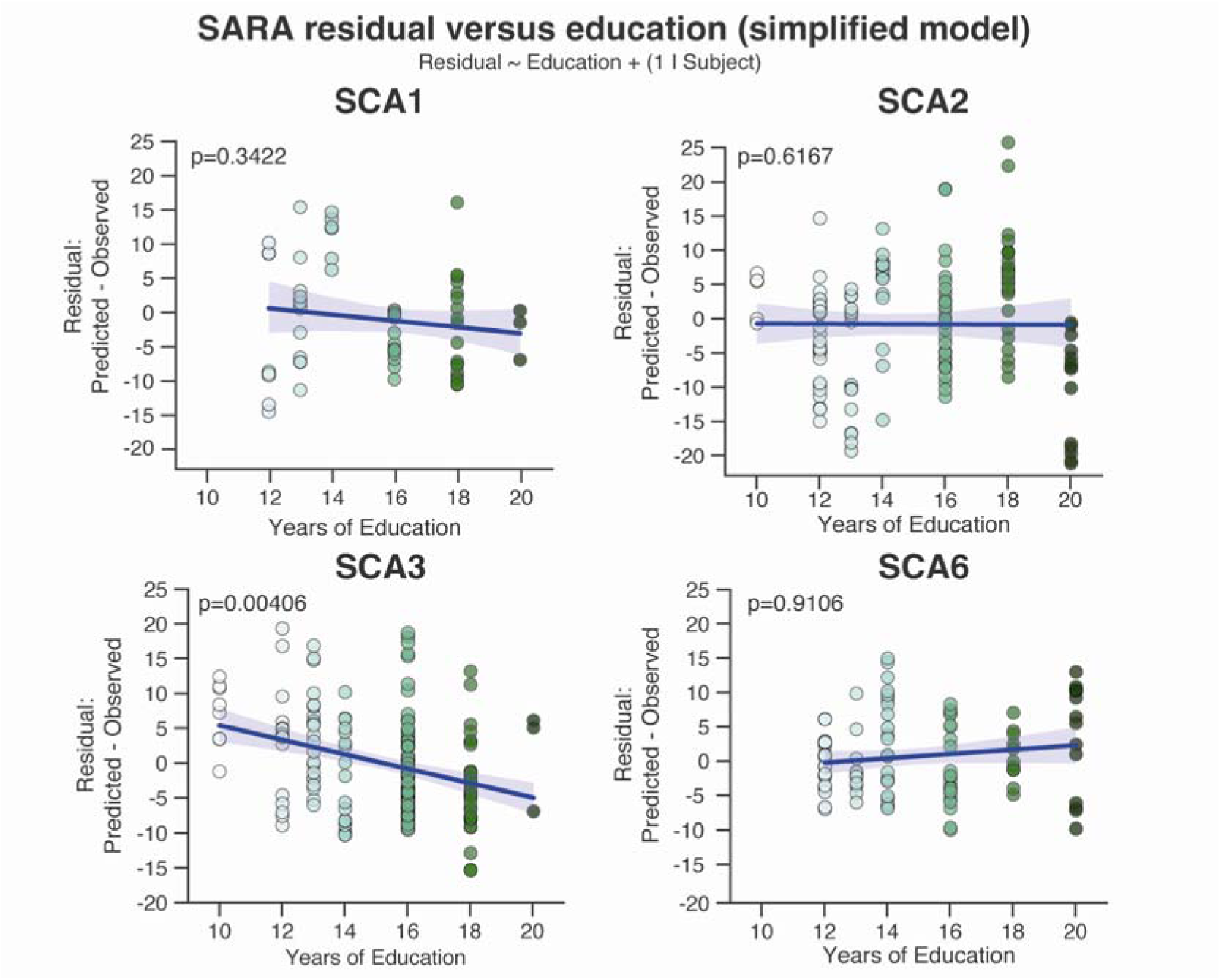
Robustness analysis 1: We built a LMM as in Figure 2, but we excluded expanded allele repeat length as an input based on collinearity with age of onset. Thus, model inputs were age of onset and duration only, and marginal residual (error) was compared to education. As in figure 2, there was a strong association between educational attainment and model error in SCA3, but not in SCA1, SCA2, or SCA6.

**Supplemental Figure 2:**
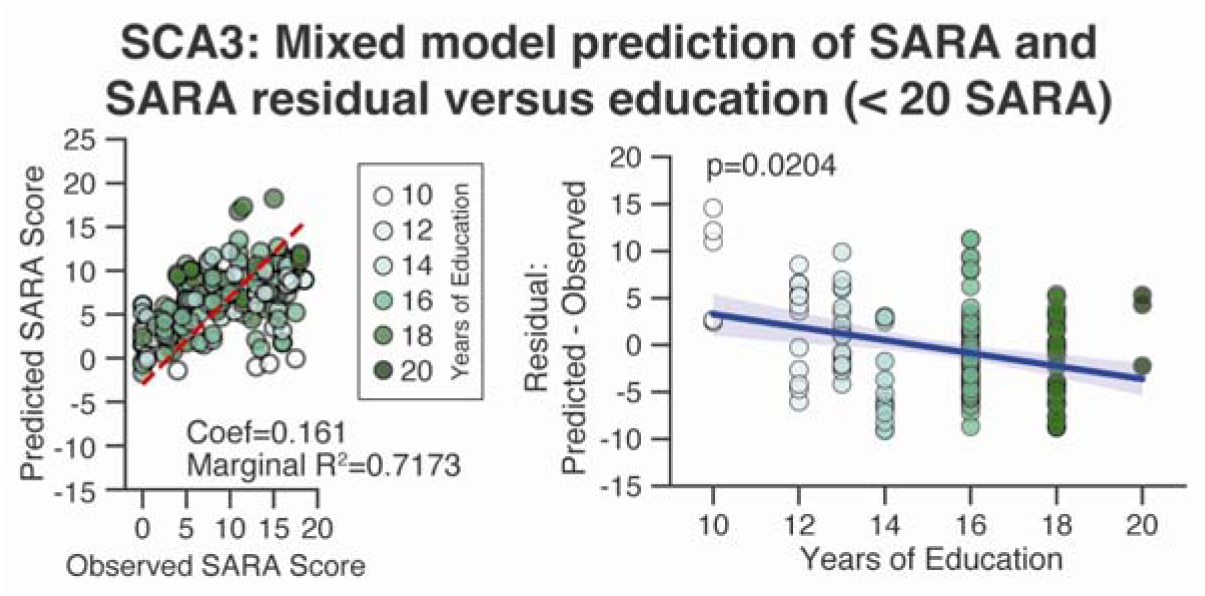
Robustness analysis 3: We built a LMM as in Figure 2, but we excluded subjects with a SARA at or above 20 to eliminate a potential confounding factor in the data, namely that patients with higher SARA scores tended to have reported less educational attainment (see Figure 2A). As with Figure 2 and other robustness analyses, primary findings held under this approach.

**Supplemental Table 2:** Coefficients (slopes) and p-values are shown for all robustness analyses.

| Robustness analysis 1: eliminating collinearity between age of onset and expanded allele repeat length by excluding expanded allele repeat length in the model (Supplemental Figure 1) |  |  |
| --- | --- | --- |
| Linear mixed-effects model |  |  |
|  | Coefficient | P-value |
| SCA1 | -0.609 | 0.342 |
| SCA2 | -0.212 | 0.617 |
| SCA3 | -0.89107 | 0.00406 |
| SCA6 | 0.039 | 0.911 |
| Robustness analysis 2: running a bootstrapping analysis instead of a leave-one-out cross validation approach |  |  |
| Ordinary least squares linear regression (ols Python function) |  |  |
|  | Coefficient | P-Value |
| SCA1 | -0.555 | 0.232 |
| SCA2 | 0.0245 | 0.94 |
| SCA3 | -0.734 | 0.00628 |
| SCA6 | -0.049 | 0.885 |
| Robustness analysis 3: evaluating only in subjects with SARA<20 to eliminate a potential confounding factor (Supplemental Figure 2) |  |  |
| Ordinary least squares multiple linear regression (ols Python function) |  |  |
|  | Coefficient | P-Value |
| SCA3 | -0.557 | 0.0204 |

## Credit Author Statement

CJA: Conceptualization, Formal analysis, Investigation, Methodology, Validation, Visualization, Writing – original draft, Writing – review and editing

DNA: Formal analysis, Investigation, Methodology, Validation, Visualization, Writing – review and editing

AK: Project administration, Data curation CRC-SCA: Data curation

VS: Data curation, Writing – review and editing SHK: Data curation, Writing – review and editing

LSR: Conceptualization, Data curation, Investigation, Methodology, Project administration, Writing – review and editing

## Acknowledgements

Clinical Research Consortium for the Study of Cerebellar Ataxia (CRC-SCA Consortium) members from 2010-February 2025 include the following:

1. Tetsuo Ashizawa, MD and Andrew Billnitzer, MD, MPH Houston Methodist Research Institute, Houston, TX, USA
2. Susan Perlman, MD Department of Neurology, University of California Los Angeles, Los Angeles, CA USA
3. Khalaf Bushara, MD Department of Neurology, University of Minnesota, Minneapolis, MN USA
4. Michael D Geschwind MD, PhD and Cameron Dietiker, MD Department of Neurology, University of California San Francisco, San Francisco, CA USA
5. Christopher M. Gomez, MD, PhD and Mahesh Padmanaban, MD Department of Neurology, University of Chicago, Chicago, IL USA
6. Sheng-Han Kuo, MD and Sandie Worley, MD Department of Neurology, Columbia University Medical Center, New York, NY USA
7. Puneet Opal, MD, PhD and Rizwan Akhtar, MD, PhD Department of Neurology, Northwestern University, Chicago, IL USA
8. Henry Paulson, MD, PhD, Sharan Srinivasan, MD, PhD, and Amy Ferng, MD Department of Neurology, University of Michigan, Ann Arbor, MI USA
9. Chiadi U. Onyike, MD Department of Psychiatry and Behavioral Sciences, Johns Hopkins University, Baltimore, MD, USA
10. Sarah Ying, MD, Liana Rosenthal, MD, PhD, and Ashley Paul, MD Department of Neurology, Johns Hopkins University, Baltimore, MD, USA
11. Jeremy Schmahmann, MD, Christopher Stephen, MD, Anoopum Gupta, MD, PhD, and Chih-Chun Lin, MD, PhD Department of Neurology, Massachusetts General Hospital, Harvard Medical School, Boston, MA USA
12. S.H. Subramony, MD and Matthew Burns, MD, PhD Department of Neurology, University of Florida, Gainesville, FL USA
13. George Wilmot, MD, PhD Department of Neurology, Emory University, Atlanta, GA USA
14. Antoine Duquette, MD, MSc Centre Hospitalier de l’Université de Montréal, University of Montreal, Montreal, QC Canada
15. Theresa Zesiewicz, MD Department of Neurology, University of South Florida, Tampa, FL USA
16. Marie Y. Davis, MD, PhD Department of Neurology, University of Washington, Seattle, WA USA
17. Ali G. Hamedani, MD, MHS and Joaquin Vizcarra Pasapera, MD Department of Neurology, Perelman School of Medicine, University of Pennsylvania, Philadelphia, PA USA
18. Vikram G. Shakkottai, MD, PhD Department of Neurology, University of Texas Southwestern Medical Center, Dallas, TX USA
19. Stefan Pulst, MD Department of Neurology, University of Utah, Salt Lake City, UT USA
20. Christian Rummey, PhD Clinical Scientist and Statistician, Basel, Switzerland
21. Lauren Moore, PhD National Ataxia Foundation, Minneapolis, MN USA

